# Multi-strain Probiotics Alter Gut Microbiota and Estrobolome Pathways in Primary Dysmenorrhea

**DOI:** 10.64898/2026.06.09.26355309

**Authors:** Siti Khadijah Kiraman, Izyan Atiqah Zakaria, Loh Sweet Yi Esther, Farisha Alia Norfuad, Nur Azurah Abdul Ghani, Hajar Fauzan Ahmad

## Abstract

**Background:** Exact cause of primary dysmenorrhoea is unknown but recent evidence uncovers a potential link between gut dysbiosis and benign gynaecological disorder via disruption of estrobolome.

**Methods:** A randomized controlled trial to investigate the effects of multi-strain oral probiotics MCP® BCMC® on primary dysmenorrhoea has been conducted. This is a secondary analysis comparing the stool microbiome in women with primary dysmenorrhoea and those without (control), and the effects of treatment with probiotics versus placebo.

**Results:** Although microbial richness and evenness were comparable between groups (alpha diversity, p > 0.05), gut microbial community composition differed significantly (Bray–Curtis PERMANOVA, p = 0.015), characterised by reduced *Bifidobacterium adolescentis* and *Blautia* and enrichment of *Faecalibacterium* in dysmenorrhoea, alongside condition-specific core taxa. Post-intervention analysis revealed significant shifts in microbial community structure between pre- and post-treatment groups (PERMANOVA, F = 2.11, R² = 0.134, p = 0.005), with probiotic supplementation inducing more consistent and directed microbiome changes than placebo, without altering alpha diversity (p > 0.05). Functional prediction showed no significant difference in overall β-glucuronidase pathway abundance (p > 0.05); however, dysmenorrhoea was associated with higher abundance of β-glucuronidase–producing taxa (MaAsLin2, q < 0.05) that were differentially modulated by probiotic treatment.

**Conclusion:** This discovery provides evidence on the microbial disruption in primary dysmenorrhoea as well as the benefit of probiotics to modulate the intestinal microbiota to improve the condition.

**What is already known on this topic:** PD is a common inflammatory gynaecological disorder that has historically been associated with hormonal imbalance and excessive prostaglandin production. Recent research indicates that the estrobolome, especially β-glucuronidase-producing bacteria may have a role in menstrual problems and inflammation. Although their impact in PD is still unclear, probiotics have also been shown to alleviate inflammatory-related disorders and modify gut flora.

**What this study adds:** Women with PD exhibited distinct gut microbial community profiles despite having comparable microbial richness and evenness to healthy controls. Dysmenorrhea was associated with reduced *Bifidobacterium adolescentis* and *Blautia*, enrichment of *Faecalibacterium*, and higher abundance of β-glucuronidase associated taxa. Multi-strains probiotic supplementation induced more consistent and directed microbiome shifts compared with placebo treatment, suggesting a modulatory effect on gut microbial composition and estrobolome-associated bacteria.

**How this study might affect research, practice or policy:** These results demonstrate the potential use of probiotics and other microbiome-targeted therapies as supplemental methods to treat primary dysmenorrhea in addition to traditional NSAID therapy. This study offers a foundation for incorporating gut microbiome and estrobolome profiling into gynaecological studies in clinical and translational research, potentially boosting the development of functional probiotics and precision therapies for illnesses related to hormones and inflammation.

## Introduction

Primary dysmenorrhea (PD) refers to painful menstruation in the absence of pelvic pathology or other organic diseases, typically occurring when the ovulatory cycle is established within the first two years after menarche in adolescent girls and young women^1–3^. It is characterized by a cramping sensation that can range widely in severity and associated symptoms, such as headaches, dizziness, fatigue, nausea, diarrhoea and mild fever^4^. The pain begins a few hours before or at the onset of menstrual bleeding. Symptoms manifestation will peak with the maximum blood flow and usually lasts up to 48 to 72 hours^5,6^. From a pathophysiological perspective, the exact cause of primary dysmenorrhea is not yet fully elucidated. However, many studies suggest that prostaglandins and other chemical mediators such as interleukins and leukotrienes, plays a significant role in the development of PD. These abnormal contractions will further result in uterine hypoxia, ischaemia due to the narrowing of blood vessels, and increases sensitivity of the nerve endings^7–9^. Non-steroidal anti-inflammatory drugs (NSAIDs) are the main treatment for PD by reducing prostaglandin production ^10^, but their limited effect on other inflammatory mediators ^11,12^ such as leukotrienes may explain why they are ineffective in some patients ^13^.

Despite the widespread use of NSAIDs and hormonal suppression as the current gold standard for PD management, these approaches primarily provide symptomatic relief without addressing the underlying inflammatory and hormonal dysregulation associated with the condition ^14,15^. Emerging evidence increasingly suggests that the gut microbiota, particularly the estrobolome, may play a significant role in PD pathophysiology through its influence on systemic inflammation and estrogen metabolism^14^. Specific gut bacteria, including *Escherichia coli, Bacteroides fragilis*, and *Streptococcus agalactiae*, produce β-glucuronidase enzymes capable of deconjugating estrogen metabolites ^16^, thereby enabling reabsorption of biologically active estrogen into circulation ^17^. Dysregulation of this process may contribute to hyper-estrogenic states and exacerbate inflammatory responses linked to PD. In this context, the limitations of NSAIDs, which do not modulate the gut microbiome or broader inflammatory networks, highlight the need for alternative therapeutic strategies. Probiotics have therefore emerged as a promising intervention due to their potential to restore microbial balance, regulate inflammatory pathways, and indirectly influence hormonal homeostasis ^18^. Although evidence supporting probiotic use in inflammatory disorders is growing ^19–21^, their role in PD remains insufficiently explored. Nevertheless, recent findings indicating that multi-strain probiotics can modulate gut microbial composition and estrobolome-associated pathways provide a compelling argument that targeting the gut microbiota may represent a novel and more mechanistic approach for PD management beyond conventional symptom suppression ^21–24^.

Therefore, in this study, we conducted a secondary analysis of data from a randomized controlled trial investigating the effects of multi-strain probiotics (MCP^®^ BCMC^®^) on women with PD. The primary aim was to compare the stool microbiome composition between women with PD and healthy controls. Additionally, this study evaluated the effects of probiotic treatment versus placebo on stool microbiota diversity and composition between across both groups. As there is limited current evidence highlighting the health benefits of probiotics as alternative treatment for dysmenorrhea, this study explored the potential of probiotics in modulating the gut microbiota. Ultimately, findings from this study will open the possibility of a natural therapeutic approach for managing PD.

## Materials and method

### Study Design

A randomized, double-blind, placebo-controlled clinical trial was conducted involving patients recruited from Universiti Kebangsaan Malaysia Medical Centre (UKMMC). The trial was registered with the U.S. National Institutes of Health Clinical Trials Registry (NCT04119011). The present study represents a secondary analysis of a previously published cohort from our group^25^, utilizing the same participant recruitment strategy and intervention framework. Participant eligibility criteria, recruitment procedures, and intervention protocols were implemented as described in the original study.

### Patients Intervention

A single centre double-blind randomised controlled trial was conducted between October and December 2019 as previously described^25^. In brief, women participants were recruited and randomized into two groups: one receiving a placebo and the other receiving oral probiotics. Participants in the probiotic group were administered a twice-daily dose for three months of a multi-strain probiotic formulation, each sachet containing 30 billion CFU of *Lactobacillus acidophilus* BCMC^®^ 12130, *Lactobacillus casei* subsp. BCMC^®^ 12313, *Lactobacillus lactis* BCMC^®^ 12451, *Bifidobacterium bifidum* BCMC^®^ 02290, *Bifidobacterium longum* BCMC^®^ 02120, and *Bifidobacterium infantis* BCMC^®^ 02129 (B-crobes Laboratory Sdn Bhd, Ipoh, Malaysia). Participants consumed the sachets either directly or mixed with room-temperature water and were instructed to store them in a dry place below 25 °C, protected from direct sunlight. The placebo group received identical sachets containing only excipients. Additionally, the participants received 250 mg of oral mefenamic acid (Ponstan) for use when needed during the study period.

Participants comprised healthy controls (HC, n = 19) and patients with primary dysmenorrhea (DYS, n = 22). The DYS group was further stratified into mefenamic acid (MA, n = 12) and co-treatment (COT, n = 17) groups, with the latter receiving both mefenamic acid and probiotics.

### Stool Sample Collection and Processing

Following recruitment, participants provided stool samples to UKMMC within 24 hours in insulated containers with cold packs, after which samples were stored at −60°C until processing. Faecal samples were thawed at 4°C, resuspended in autoclaved Milli-Q water (1:2, w/v), homogenized for 1 min, and aliquoted into 2 mL vials. DNA was extracted from 200 mg of fecal material using the QIAamp PowerFecal Pro DNA Kit (QIAGEN, Germany), incorporating bead-beating with a FastPrep system to improve cell lysis. The quality and concentration of extracted DNA were assessed using a NanoDrop 1000 Spectrophotometer (Thermo Fisher Scientific, DE, USA), and samples were stored at −20[°C until further analysis.

### DNA Library Preparation, Amplicon Sequencing and Raw Microbial Genomics Data Analysis

The gut prokaryotic community composition was assessed through 16S rRNA gene amplicon sequencing targeting the V3 region, using the Illumina NextSeq 500 platform. Brifly, the workflow included an initial amplification (1st PCR), indexing/barcoding (2nd PCR), purification of the amplicon library, and sequencing, all conducted according to the protocol described previously^26^.

Raw paired-end reads were quality-filtered, adapter-trimmed, and merged using fastp v0.21^27^. Primer sequences located at the 5′ and 3′ ends of the merged reads were subsequently removed using cutadapt v1.18^28^. The resulting reads were then denoised with DADA2 within the QIIME2 v2021.4 pipeline^29^. Taxonomic classification of Amplicon Sequence Variants (ASVs) was performed using the q2-feature-classifier plugin^30^, trained on the V3-trimmed region of the 16S rRNA reference sequences from the latest GTDB 16S rRNA V3 region database (r202 release). ASVs classified only at the kingdom level (Bacteria) were further examined using BLAST against the NCBI nucleotide database.

### Comprehensive Statistical Analysis of Microbiome Data, Biomarkers and Functional Analyses

Non-microbial and human-derived ASVs were excluded prior to downstream analysis, and curated ASV and taxonomic tables from QIIME2 were prepared for MicrobiomeAnalyst2^31^. MicrobiomeAnalyst2 was used to profile bacterial communities across study groups, with low-abundance and low-variability features removed before data normalization using cumulative sum scaling (CSS). Alpha diversity was assessed using Chao 1, Shannon, and Observed species indices, while beta diversity was evaluated with Bray–Curtis dissimilarity metrics, and statistical significance was set at p < 0.05. Core microbiome analysis was performed with a 0.01% abundance threshold and a 50% sample prevalence cut-off. Functional predictions of 500 most abundant taxa were generated using PICRUSt2^32^ using the Kyoto Encyclopedia of Genes and Genomes (KEGG) database. Enriched pathways were visualized in STAMP v2.1.3^33^, with group differences assessed by one-way ANOVA followed by Tukey–Kramer post hoc comparisons at a 95% confidence level. Effect sizes were calculated using eta-squared (η²), and multiple testing was controlled with the Benjamini–Hochberg false discovery rate (FDR) correction. Additionally, Microbiome Multivariate Association with Linear Models (MaAsLin2) v1.22.0^34^ was applied to identify potential microbial biomarkers.

## Results

### Gut microbiome profiling in primary dysmenorrhea

The relative abundance of bacterial taxa was compared between the HC and DYS groups at the species level (Fig 1a). Across all samples, the gut microbiota was dominated by *Blautia* sp., *Faecalibacterium* sp., and *Bifidobacterium adolescentis*. The DYS group showed a lower relative abundance of *B. adolescentis* and *Blautia* sp. compared to the HC group, whereas *Faecalibacterium* sp. was more abundant in the DYS group.

**Fig 1.**
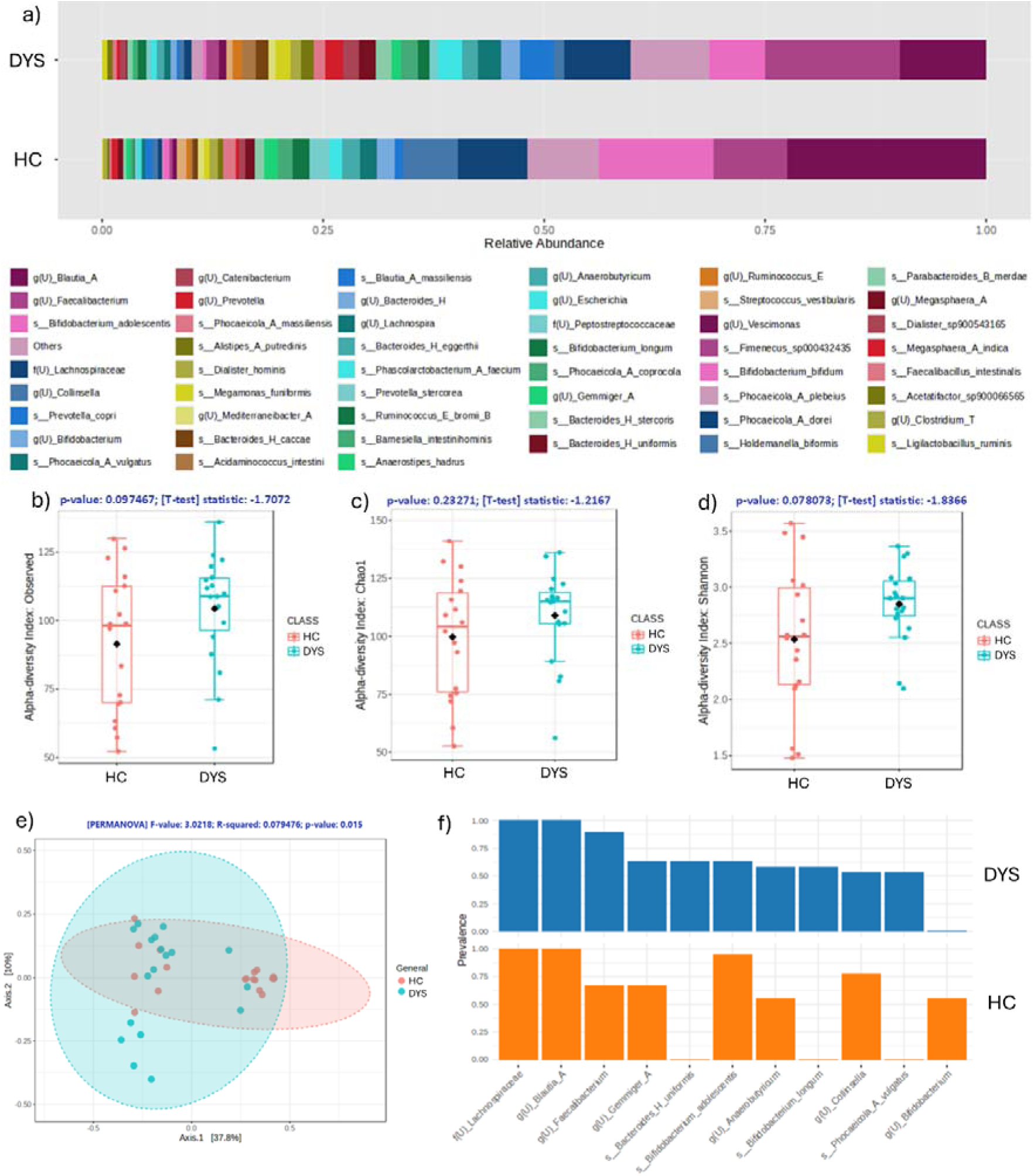
(a) Relative abundance of bacterial community in gut of DYS and HC individuals in the form of bar plots illustrating the taxa summary of merged samples at species level. The box plot illustrates the (b) (c) (d) alpha diversity; and (e) 2D PCoA plot for beta diversity analysis of bacterial communities of DYS and HC groups; (f) Core microbiome analysis of DYS and HC in identifying predominant bacterial taxa.

Alpha diversity between the HC and DYS groups was evaluated, and show no significant differences in microbial richness or evenness between the two groups (Fig 1b-1d). However, beta diversity analysis performed by Bray-Curtis dissimilarity revealed a significant difference in overall microbial community composition between HC and DYS groups (PERMANOVA, *p*-value = 0.015) (Fig 1e).

Core microbiome revealed features that are unique to the groups as well as stable commensals presence throughout the gut microbiome of HC and DYS (Fig 1f). Throughout the two groups, the presence of Lachnospiraceae, *Blautia*, *Faecalibacterium* and *Bifidobacterium adolescentis* is widely available between two groups. In contrast, the genus *Bifidobacterium* was detected only in the HC group and not in the DYS group, indicating its potential role as a core taxon in healthy controls. Conversely, *Bacteroides uniformis*, *Phocaeicola vulgatus*, and *Bifidobacterium longum* were observed exclusively in the DYS group.

### Comparative Effectiveness of Intervention treatments for dysmenorrhea

Alpha diversity indices show no significant in the differences between richness and evenness of the sample (*p*-value > 0.05) (Fig 2a-c). The PCoA plot based on Bray-Curtis dissimilarity visualised partial separation among the four groups that is pre-MA/COT and post-MA/COT (PERMANOVA, *F*-value = 2.1102, R^2^ = 0.1338, *p*-value = 0.005) (Fig 2d).

**Fig 2.**
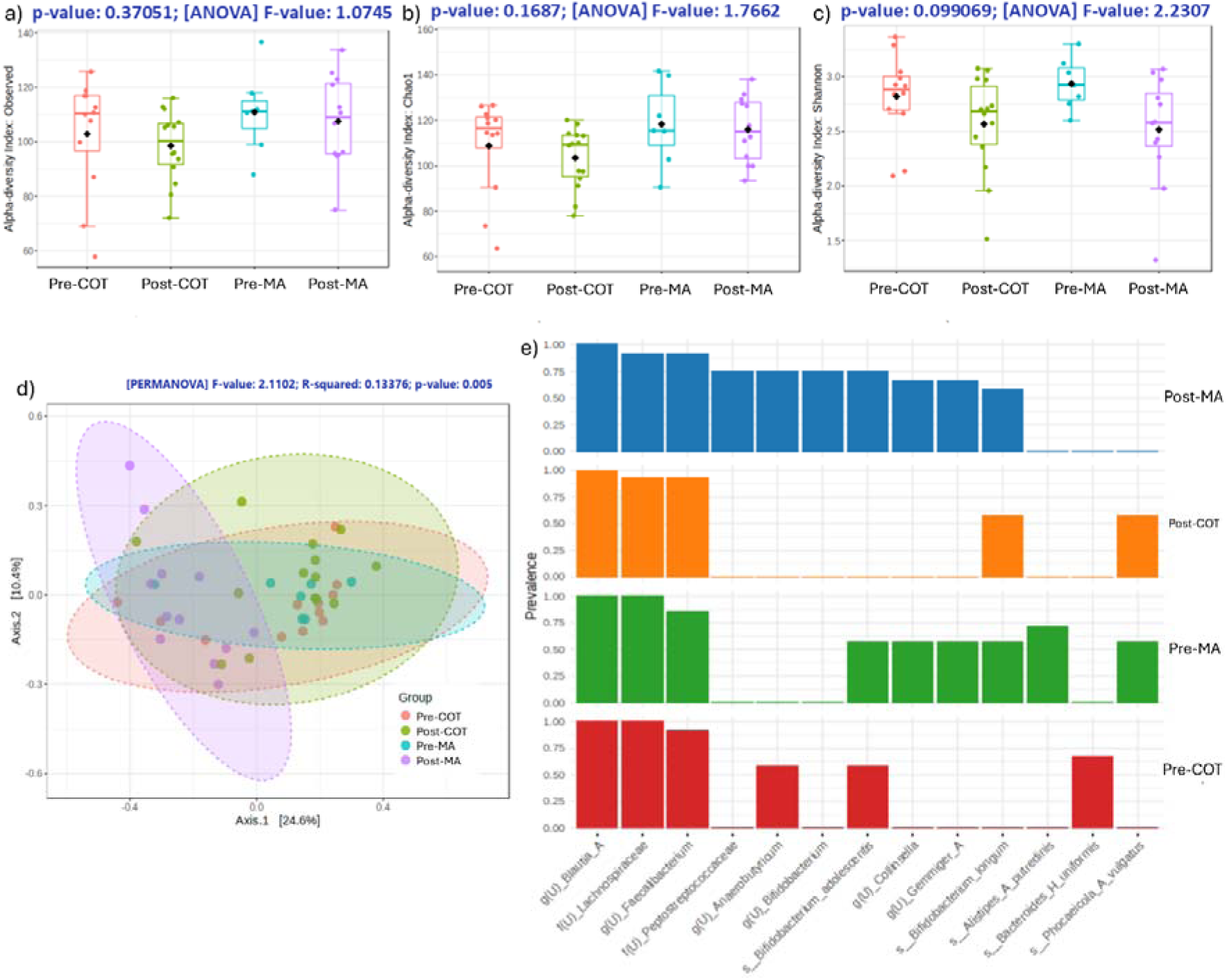
(a) (b) (c) The box plot illustrates the alpha diversity; and (d) 2D PCoA plot for beta diversity analysis of bacterial communities of pre- and post-intervention groups (e) Core microbiome analysis of pre- and post-interventions in identifying predominant bacterial taxa.

For the core microbiome, the COT intervention induced a clear shift in the gut microbiome. Several species present before COT, such as *Anaerobutyricum* and *Bifidobacterium adolescentis*, disappeared afterward, while *Bifidobacterium longum* and *Phocaeicola vulgatus* emerged. In the MA (placebo) group, microbial changes were more random and likely reflected natural variation rather than an intervention effect. *Alistipes putredinis* and *Phocaeicola vulgatus* were lost post-MA, whereas Peptostreptococcaceae, *Anaerobutyricum*, and *Bifidobacterium* appeared (Figure 2e).

### Decoding the ***β***-Glucuronidase Axis: Discriminant Microbial Determinants Identified by PICRUSt2 and MaAsLin2

PICRUSt2 and MaAsLin2 analyses revealed that although the interventions did not significantly alter overall β-glucuronidase activity, clear differences in estrobolome-associated microbial composition were observed between healthy controls and dysmenorrhea subjects. PCA revealed overall differences in microbial composition among pre- and post-intervention groups where the first three principal components explained 52.6%, 13.7%, and 10.3% of the total variance, respectively (Figure 3a). Several β-glucuronidase–producing taxa, including members of the *Bacteroides* and *Prevotella* genera, were consistently less abundant in healthy controls, supporting their potential involvement in PD-associated hormonal and inflammatory dysregulation. Following intervention, *Bacteroides uniformis* and *Prevotella copri* decreased in both treatment groups, suggesting a shared microbial response, while *Bacteroides eggerthii*, *Bacteroides* H, and *Alistipes putredinis* decreased only in the MA group but remained relatively stable in the COT group (Fig 3c-h), which parallel to previous findings (Table 1). These findings suggest that the two interventions may differentially influence estrobolome-associated taxa despite having minimal impact on overall predicted β-glucuronidase function.

**Fig 3.**
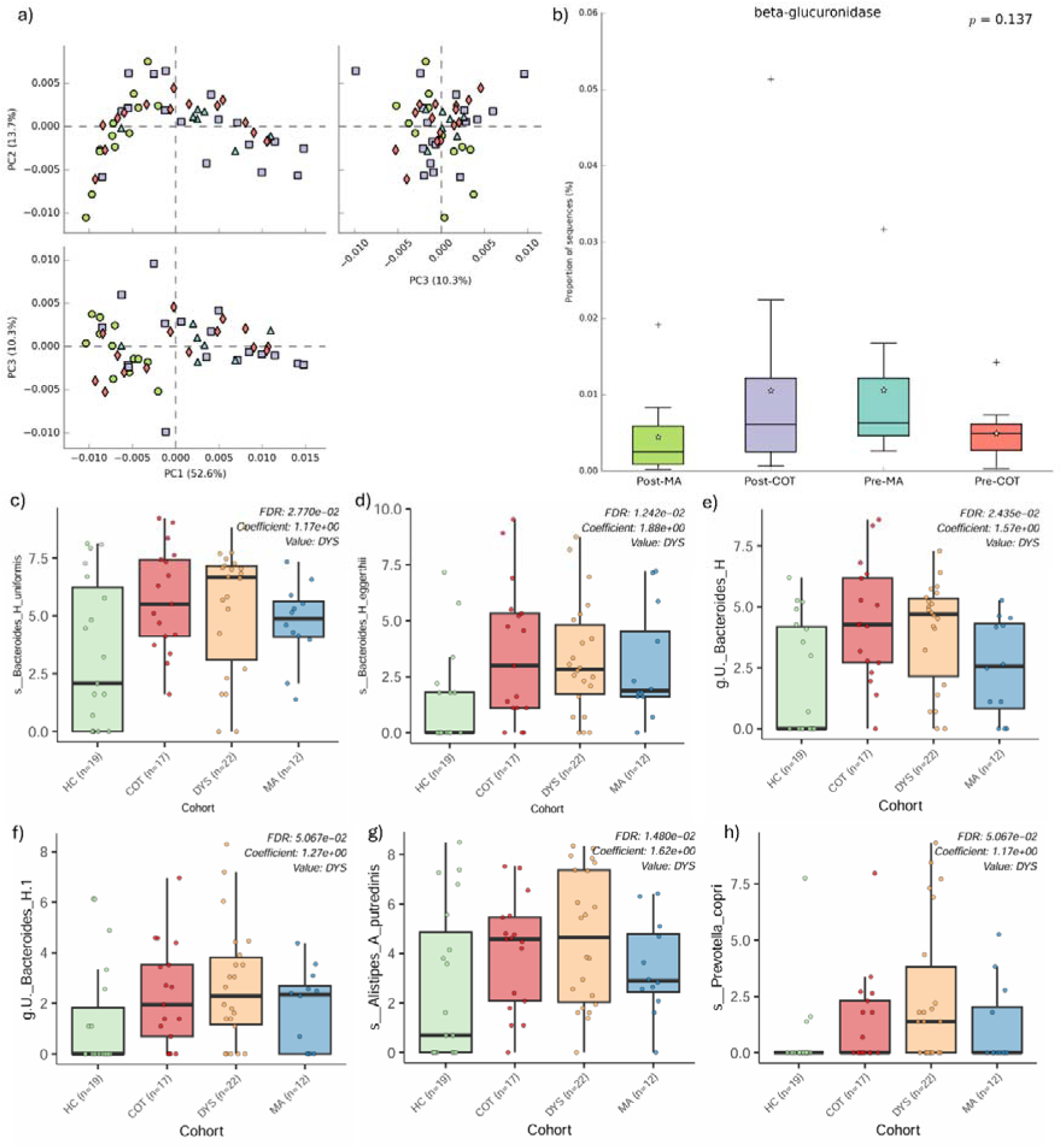
Statistical Analysis of Metabolic Profiles (STAMP) analysis (a) Principal Component Analysis (PCA) plot; (b) Comparative analysis of β-glucuronidase metabolic pathways between pre- and post-interventions; (c) (d) (e) (f) (g) (h) Potential β-glucuronidase producers.

**Table 1.** List of bacterial taxa potentially to be β-glucuronidase contributors.

| No. | β-glucuronidase producers | Coefficient value | Standard error | p-value | q-value | Literature |
| --- | --- | --- | --- | --- | --- | --- |
| 1 | *Bacteroides uniformis* | 1.1718 | 0.3426 | 1.0780e-03 | 2.7697e-02 | <sup>35</sup> |
| 2 | *Bacteroides eggerthii* | 1.8753 | 0.4876 | 2.7294e-04 | 1.2422e-02 | <sup>36</sup> |
| 3 | *Bacteroides H* | 1.5688 | 0.4498 | 8.7118e-04 | 2.4347e-02 | <sup>36,37</sup> |
| 4 | *Bacteroides H.1* | 1.2727 | 0.4254 | 3.8949e-03 | 5.0671e-02 | <sup>36,37</sup> |
| 5 | *Alistipes putredinis* | 1.6161 | 0.4498 | 8.7118e-04 | 2.4347e-02 | <sup>36</sup> |
| 6 | *Prevotella copri* | 1.1706 | 0.3905 | 3.8281e-03 | 5.0671e-02 | <sup>36</sup> |

### Mapping Microbial Fingerprints Across Study Cohorts

This section employed MaAsLin2 to identify microbial biomarkers across the study cohorts (Fig 4). *Evtepia gabavorous*, a bacterium reported to utilize γ-aminobutyric acid (GABA) for growth^38^, increased in abundance following both interventions, despite low baseline levels in the HC and DYS groups, indicating an intervention-associated effect. *Alistipes putredinis* was more abundant in the DYS group than in the HC group and decreased following the MA intervention, with little to no change observed after COT, suggesting greater efficacy of MA in reducing this taxon. *Negativibacillus massilinensis* showed higher relative abundance in the DYS group compared with the HC group. The two interventions yielded contrasting effects, the MA intervention resulted in minimal reduction, whereas the COT intervention led to near-complete depletion, suggesting a potential role of the probiotic in reducing this taxon.

**Fig 4.**
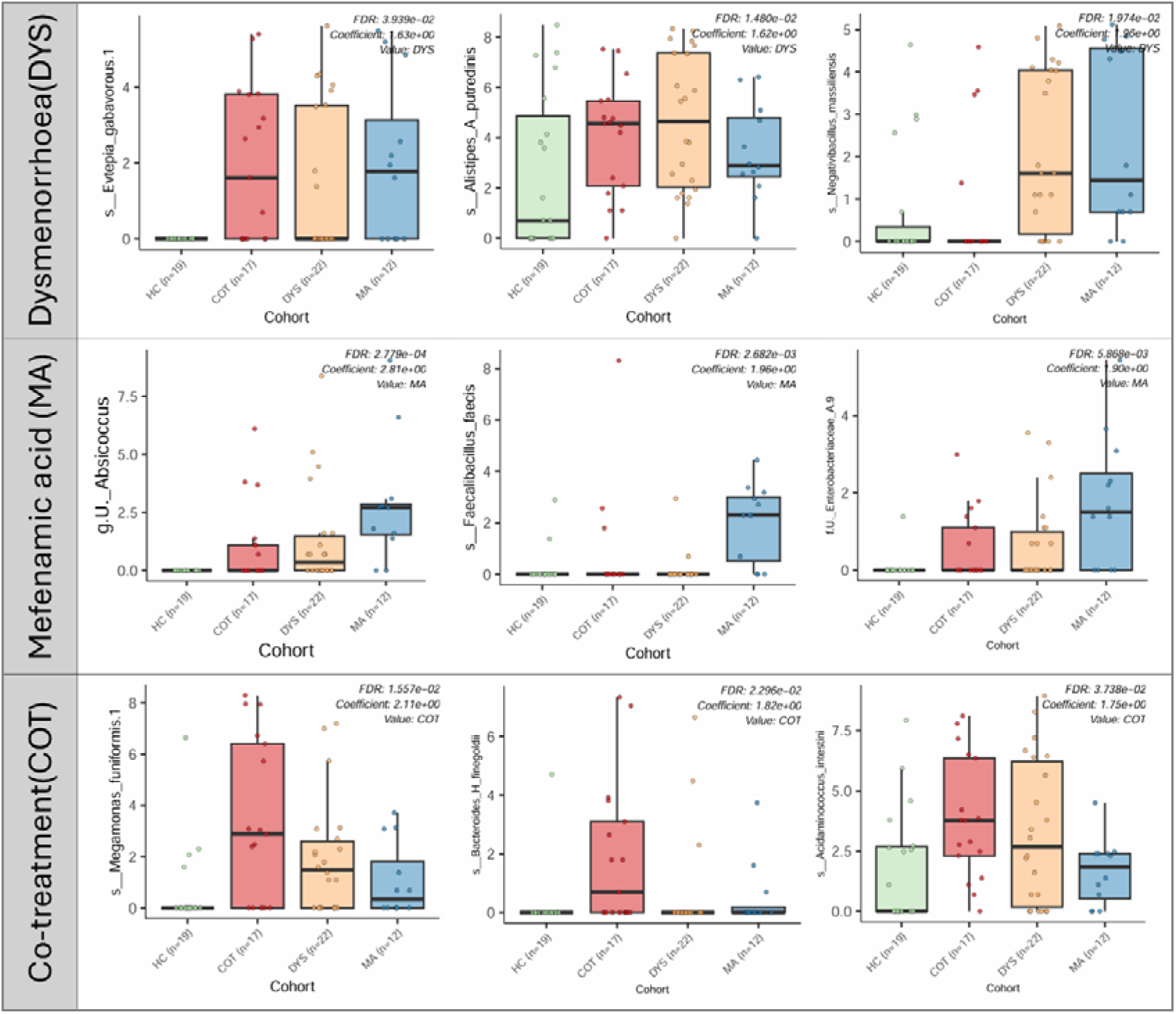
Each cohort potential biomarkers identified by MaAsLin2 adjusting for cohort with HC as baseline

Focusing on biomarkers associated with the MA intervention, these biomarkers exhibited low relative abundance in the HC group, suggesting that they are primarily associated with the DYS condition and/or intervention effects. *Absicoccus* and *Enterobacteriaceae* A.9 showed higher relative abundance in the MA group compared with the DYS group and lower abundance in the COT group, indicating that the MA intervention may promote the enrichment of these taxa. *Faecalibacillus faecis* was uniquely detected in the MA group, with little to no presence in the HC, DYS, and COT groups, suggesting that its occurrence may be specifically associated with the MA intervention or characteristics unique to the MA-treated participants.

The COT intervention biomarkers exhibited higher relative abundance in the COT group, while remaining at very low abundance in the HC group. *Meganomonas funiformis* showed increased relative abundance following the COT intervention compared with the DYS group, whereas a decrease was observed in the MA group. *Bacteroides finegoldii* was detected predominantly in the COT group and showed minimal presence in other groups. Additionally, *Acidaminococcus intestini* increased in abundance following the COT intervention relative to the DYS group, while a decrease was observed following the MA intervention.

## Discussion

Beta diversity analysis revealed significant differences in microbial community composition between the HC and DYS groups (*p* = 0.015), suggesting that PD is associated with shifts in specific gut microbial taxa rather than overall microbial richness. In contrast, alpha diversity showed no significant differences between groups (*p* > 0.05), although slightly higher diversity was observed in the DYS group. This finding argues that greater microbial diversity does not necessarily indicate a healthier gut microbiome, as increased diversity may also reflect altered gut transit, dysregulated fermentation ^39^, or elevated serum oestrogen levels ^40^.

The top three taxa by relative abundance in both the HC and DYS groups were *Blautia* spp., *Faecalibacterium* spp., and *Bifidobacterium adolescentis*. *Blautia* spp. are key commensals in the gut and are known to contribute to intestinal homeostasis through the production of metabolites that support mucosal integrity and protect the intestinal epithelium from pathogenic invasion^41^. The reduced abundance of *Faecalibacterium* spp. has been consistently associated with inflammatory conditions^42^. Notably, *B. adolescentis* has been reported in previous studies to be more prevalent in females, which is consistent with the microbial composition observed in our cohort^43^. In parallel, core microbiome analysis supported these findings, demonstrating that these three taxa were present in at least 70% of samples at an abundance threshold of 0.03%, indicating their classification as core members of the gut microbiome in this study (Fig 1f).

While humans perform glucuronidation in the liver to detoxify xenobiotics, inactivate drugs, and facilitate their excretion via bile and urine^44^, gut microbes expressing β-glucuronidase can reverse this process by deconjugating glucuronidated compounds and regenerating their active forms within the intestine^45^. The presence of microbial taxa harbouring β-glucuronidase homologs therefore represents a critical control point for chemical reactivation in the gut. In individuals using NSAIDs, such as mefenamic acid for primary dysmenorrhoea, microbial β-glucuronidase–mediated deconjugation can lead to drug reactivation in the colon, thereby increasing the risk of intestinal toxicity^46^. In addition, β-glucuronidase is a central component of the oestrobolome^47^, and elevated enzyme activity has been associated with disturbance in circulating oestrogen levels and a higher risk of oestrogen-dependent conditions, including gynaecological cancers and menopausal syndrome^47^.

Elucidating potential β-glucuronidase–producing taxa may facilitate the identification of oestrobolome-related biomarkers^47^. Previous study has demonstrated that gut microbial β-glucuronidase enzymes can deconjugate inactive oestrogen glucuronides, thereby reactivate oestrogens and enable their reabsorption into blood circulation^48^. Although PD is not traditionally classified as an oestrogen-dependent disorder, oestrogen plays a critical role in its pathophysiology by upregulating cyclooxygenase-2 (COX-2) expression in endometrial tissue^49^. Excessive prostaglandin production, a hallmark of dysmenorrhea symptoms^13^.

Using MaAsLin2, several potential biomarkers were identified, with supporting evidence from the literature suggesting their association with β-glucuronidase activity. Among these taxa, *Bacteroides uniformis,* present to be in high abundance compared to other cohorts in Fig 3c, has been reported to produce multiple β-glucuronidase enzymes in the gut^35^, whereas other taxa, including *Bacteroides eggerthii*, *Bacteroides* spp., *Alistipes putredinis*, and *Prevotella copri*, have primarily been reported to encode putative β-glucuronidase homologs^36,37^. For these latter taxa, further experimental investigation is required to confirm enzyme expression and functional activity. Notably, major bacterial phyla commonly associated with β-glucuronidase activity include Bacteroidetes, Firmicutes, Proteobacteria, and Actinobacteria, with several members of the genus *Bacteroides* reported to encode β-glucuronidase homologs^16^.

Compared with the HC group, all taxa exhibited higher relative abundance in the DYS group. However, the presence of these taxa does not necessarily indicate a direct contribution to β-glucuronidase activity in individuals with PD, as these microorganisms may exert additional biological functions beyond enzyme production. The excessive β-glucuronidase–producing bacteria may disrupt hormonal balance, highlighting the potential of microbiome-based strategies for early detection and disease prevention^50^.

Alpha diversity showed similar microbial richness between the MA and COT groups, while beta diversity revealed significant differences in microbial composition before and after intervention. Although the gut microbiota remained relatively stable after COT treatment, the intervention produced a clearer and more consistent shift in microbial community structure compared to MA.These findings may explain the reduced analgesic use and improved mental health scores observed in the clinical outcomes reported by^25^.

In contrast, the MA group exhibited greater dispersion, indicating higher inter-individual variability following treatment. This pattern suggests that MA alone elicited heterogeneous microbial responses rather than a consistent, intervention-driven shift in community composition. Conversely, the COT intervention appeared to preserve a degree of compositional stability despite microbial changes. In essence, these observations suggest that the MA intervention induced greater microbial variation, while the COT intervention may have contributed to maintaining microbial stability through probiotic action. Given that *Bifidobacterium* is known to support the growth and persistence of other gut commensal microbes by cross-feeding^51^, the inclusion of probiotics alongside mefenamic acid may contribute to maintaining overall microbial community structure.

Next, this section discusses cohort-specific gut microbiome biomarkers identified by MaAsLin2 and their potential roles in inflammation and endocrine-related conditions. In the DYS cohort, *Evterpia gabavorous*, *Alistipes putredinis* and *Negativibavillus massiliensis* are the top three of biomarkers associated with this cohort. *E. gabavorous* is species known to consume GABA as part of its growth^38^, indicating symbiosis with other GABA-producer such as *Bacteroides* genus^52^. Given the association of primary dysmenorrhea with anxiety and depression^53^, the enrichment of *Bacteroides* (Fig 3) and *E. gabavorous* (Fig 4) in the

DYS cohort may reflect microbial involvement in GABA-related metabolic processes, which could influence gut–brain signalling and mental state. As shown in Fig 4, *E. gabavorous* differed in abundance between the HC and DYS cohorts and increased following the intervention. This enrichment may reflect changes in the gut environment that favour the persistence of this species. In parallel, Fig 3 shows that *Bacteroides* species were abundant across most cohorts except HC, suggesting cohort-specific microbial configurations that may shape a GABA-centred microbial network.

*A. putredinis*, a core member of the human gut microbiome, is typically associated with healthy individuals^54^ and showed a protective or homeostatic role^55,56^. Its enrichment in the DYS cohort may reflect a compensatory response to inflammatory changes. However, as noted previously, *A. putredinis* also harbours genes encoding β-glucuronidase, which may influence oestrogen-related pathways relevant to dysmenorrhea. Taken together, these observations suggest that its association with lower inflammation reflects a nuanced and context-dependent role in this cohort.

In the post-MA cohort, minority biomarkers included *Absicoccus* sp., and Enterobacteriaceae, all detected at low abundance in HC, although their biological relevance remains unclear due to limited functional characterization and taxonomic resolution.

In contrast, *Faecalibacillus faecis*, as shown in Fig 4, appears to be uniquely enriched in the MA cohort. This species is a human gut–associated bacterium belonging to the family Erysipelotrichaceae^57^. Notably, genomic analyses have reported negative β-glucuronidase activity for *F. faecis*^57^, suggesting that its role in the MA cohort may reflect cohort-specific microbial characteristics rather than mechanisms directly associated with primary dysmenorrhea.

Focusing on the COT intervention, which combined mefenamic acid administration with HEXBIO^®^ probiotic culture supplementation, *Megamonas funiformis*, *Bacteroides finegoldii*, and *Acidaminococcus intestini* were identified as the top three microbial biomarkers. *Megamonas funiformis* is considered a metabolic indicator taxon, as it has been widely reported to be diet-responsive, particularly to complex carbohydrates and dietary fibres such as inulin *via* fermentation^58,59^. Previous study suggests that *M. funiformis* may alleviate metabolic dysfunction–associated fatty liver disease through propionic acid production, which inhibits *de novo* lipogenesis and promotes β-oxidation, supporting its potential as a probiotic candidate^58^. Its enrichment in the COT cohort likely reflects altered substrate availability and enhanced microbial metabolic activity rather than a direct pathogenic role. These findings suggest that supplementation with HEXBIO^®^ probiotic cultures may have increased the availability of fermentable substrates that favour the growth and metabolic activity of *M. funiformis*.

*Bacteroides finegoldii* appear unique to the COT cohort, associated for its potential as psychobiotic^60^. Although the roles of this bacteria is still understudied^61^, the presence of *B. finegoldii* exclusively in the COT cohort suggests a synergistic effect of probiotic supplementation and NSAID treatment on gut microbial ecology of patients with PD.

Interestingly, in contrast to the other two COT-associated biomarkers, *Acidaminococcus intestini* is a diet-responsive taxon primarily associated with amino acid rather than carbohydrate metabolism^62^. To date, there are no reports implicating *A. intestini* in bacteremia or pathogenicity. Instead, this species predominantly ferments amino acids specifically glutamic acid to produce short-chain fatty acids^62^, rendering it highly responsive to protein availability and microbial cross-feeding within the gut ecosystem.

Overall, the COT intervention suggests that the observed microbial changes are largely driven by diet-responsive taxa rather than by microbes with direct mechanistic involvement in disease processes. Probiotic supplementation may have enhanced microbial responsiveness to available dietary substrates, thereby promoting the enrichment of metabolically active taxa. The identified biomarkers therefore appear to play a supportive or adaptive role within the gut ecosystem rather than acting as primary drivers of pathology. Importantly, the combined intervention did not indicate any concerning microbial shifts but instead reflects a transition toward a more metabolically active and functionally engaged fermentation network. Importantly, no enrichment of taxa associated with inflammation, pathogenicity, or dysbiosis was observed, suggesting that the combined intervention did not induce adverse microbial shifts.

This study has several limitations. First, the sample size for each cohort was relatively small. Second, the findings are based on 16S rRNA amplicon sequencing rather than metagenomic or metatranscriptomic approaches, which restricts functional resolution. Consequently, the analyses focused on taxonomic biomarkers rather than direct assessment of gene expression or metabolic activity. As gene presence does not necessarily equate to functional expression, further studies incorporating functional omics and experimental validation are warranted.

## Conclusion

This study shows that although microbial diversity was comparable between the HC and DYS groups, their community compositions differed significantly, suggesting that specific microbial shifts rather than overall diversity are associated with primary dysmenorrhea. The identification of β-glucuronidase–associated taxa further highlight a potential link between gut microbiota, oestrogen metabolism, and disease pathophysiology. In addition, while both MA and COT interventions exhibited similar effects on microbial richness, they produced distinct compositional changes. The COT intervention, particularly with probiotic supplementation, appeared to promote greater microbial stability and metabolic activity, whereas MA alone resulted in more variable responses. Overall, these findings underscore the potential role of the gut microbiome in primary dysmenorrhea and suggest that microbiome-targeted strategies may offer therapeutic value. However, further studies with larger sample sizes and functional validation are required to confirm these associations.

## Author Contributions Statement

I.A.Z and N.A.A.G conceived and designed the study. I.A.Z, L.S.Y.E and F.A.N performed the experiments and analyzed the data together with S.K.K and H.F.A. I.A.Z and S.K.K. drafted the manuscript with input from N.A.A.G, and H.F.A. All authors reviewed and approved the final version of the manuscript for publication.

## Data Availability

The sequences for the 16S rRNA data were deposited in the NCBI database and registered under BioProject PRJNA1399960.

https://www.ncbi.nlm.nih.gov/bioproject/PRJNA1399960/

## Acknowledgements

The authors would like to thank all the clinical and supporting staff of the Department of Obstetrics and Gynecology, Hospital Canselor Tunku Muhriz, for their support and involvement throughout the study. We would like to acknowledge B-Crobes Laboratory Sdn. Bhd. for a 3-month supply of placebo and probiotic sachets.

## Patient and public involvement

Patients or the public were involved in the design, or conduct, or reporting, or dissemination plans of our research.

## Consent for publication

All participants provided consent for the publication of data generated from this study.

## Ethical approval statement

Human ethical approval and consent to participate This study was conducted in accordance with the principles outlined in the Declaration of Helsinki. Ethical approval was obtained from the Institutional Ethics Committee of the National University of Malaysia (FF-2018-204; approved on 5 June 2018), and the study was registered at ClinicalTrials.gov (NCT04119011). Written informed consent was obtained from all participants prior to their inclusion in the study.

## Animal ethical approval

This study does not involve animal subjects.

